# Intrathecal Dexmedetomidine as an adjunct for Labour Analgesia: A Systematic review

**DOI:** 10.1101/2025.07.11.25331341

**Authors:** Adanech Shifarew Legasse, Fikadu Tilahun, Yesuf Ahmed, Bekele Buli, Addisu Mossie, Aschalew Besha, Selam Tamiru

## Abstract

**Introduction:** Labor pain is one of the most painful experiences that women encounter during child birth that affect fetal and maternal wellbeing. Therefore providing labor analgesia is an essential part of maternity care for laboring mother. Intrathecal dexmedetomidine has emerged as a promising option for labor analgesia adjunct with probable less side effects than opioids in recent years. The aim of this review is to systematically review the current literature on the efficacy, safety and side effect profile of intrathecal Dexmedetomidine as a non-opioid option adjunct for labour epidural analgesia.

**Methods:** This systematic literature review is reported using the Preferred Reporting of Systematic Review and Meta-Analysis protocol. The search aimed to identify articles published in the English language that discussed the efficacy of intrathecal Dexmedetomidine for combined spinal epidural or intrathecal labor analgesia that published before February 2023. To explore relevant and related articles related to our research question (PICO question), we utilized various search engines including Google Scholar, PubMed/Medline, the Cochrane Library, and conducted manual searches.

**Result:** The search strategy retrieved 38 articles from various electronic databases and after removing duplicates by Endnote 20 version and 32 articles were selected for screening; finally, nine articles were included for systematic review and critical appraisal.

**Conclusion:** This review indicates that compared to opioids intrathecal administration of Dexmedetomidine for labor analgesia provides potent analgesic efficacy, improving the duration, intensity, and quality of the block, reducing the need for opioids and minimizing opioid related adverse effects for both mothers and neonates.

## Introduction

Labor pain is one of the most painful experiences that women encounter during child birth(1). Even though for many women giving birth is a normal experience, it can have long-term morbidity on the fetus and the mother in the form of postpartum depression, persistent pain, and developmental problems (1). Therefore providing labor analgesia is an essential part of maternity care for laboring mother. (2)

Intrathecal dexmedetomidine (Dex) is a technique that involves the administration of dexmedetomidine, a selective alpha-2 adrenergic agonist, directly into the intrathecal space to provide pain relief (3, 4). Dexmedetomidine acts by binding to alpha-2 adrenergic receptors in the central nervous system, resulting in sedation, analgesia, opioid-sparing properties, sympatholytic and anxiety-relieving (4, 5). When administered intrathecally, it targets the pain transmission pathways in the spinal cord, providing effective pain relief and it has been used successfully as adjuvant agent in regional anesthesia to prolong the duration of motor and sensory blockade, lessen the toxicity associated with local anesthetic administration, and accelerate the onset of surgical block. (3, 6)

Neuraxial labor analgesia is the most complete and effective method of labor analgesia. (7) Epidural labor analgesia is the gold standard and is a very promising, safe and effective method of providing pain relief in obstetric analgesia. (8) Whereas, Combined spinal epidural (CSE) techniques provide a significantly more rapid onset of analgesia, potentially making this technique beneficial in parturients whose labor is rapidly progressing. (9) Intrathecal labor analgesia is simple, easy, and effective method for painless and safe delivery; especially single shot intrathecal (spinal) labor analgesia is promising neuraxial labor analgesia technique for resource limited settings.

However, higher doses of neuraxial local anesthetics administered in women undergoing the birthing process can impair motor function that can result in a higher rate of instrumental delivery and cesarean sections. (10) Neuraxial local anesthetics and opioids appear to act synergistically to provide neuraxial analgesia when used as low dose local anesthetics and opioid as adjuvant; and thus these reduces side effects related with high dose local anesthetics. (7) However, opioids are associated with undesirable maternal and neonatal side effects such as pruritus, nausea, vomiting, respiratory depression and neonatal sedation. (11)

Intrathecal dexmedetomidine as adjunct to local anesthetics has emerged as a promising option for labor analgesia with probable less side effects than opioids in recent years. The aim of this review is to systematically review the current literature on the role; efficacy, safety and side effect profile of intrathecal Dexmedetomidine as a non-opioid option adjunct for labour epidural analgesia. This systematic review can contribute to providing evidence-based decision-making, improving patient care, and guide further research in advancing better labor analgesia.

## Methods

This systematic literature review is reported using the Preferred Reporting of Systematic Review and Meta-Analysis (PRISMA) protocol. (12)

### Eligibility criteria

This review included all randomized controlled trials published in the English language that investigated the effects of intrathecal dexmedetomidine for labor analgesia and has full text. However, we excluded studies conducted on populations other than laboring mothers, as well as articles that examined the effects of dexmedetomidine through routes of administration other than intrathecal, studies that explored peripheral nerve block without labor pain management, studies with abstracts only, and studies published in languages other than English.

### Information sources

We utilized various search engines including Google Scholar, PubMed/Medline, the Cochrane Library, and manual search was searched studies done before February 2023. In addition, the reference lists of included articles were also hand-searched. The search employed keywords, phrases, and specific subject headings.

### Search strategy

A comprehensive search was conducted with the aim to identify articles published in the English language that discussed the efficacy of intrathecal dexmedetomidine for epidural labor analgesia. To explore relevant and related articles related to our research question (PICO question), we utilized various search engines including Google Scholar, PubMed/Medline, the Cochrane Library, and conducted manual searches. The search employed keywords, phrases, and specific subject headings.

The search strategies were developed using different electronic database strategies, and each keyword was connected using the Boolean operators “AND” or “OR” (Intrathecal OR spinal OR combined spinal epidural AND dexmedetomidine OR alpha-2 agonist AND labor AND analgesia OR pain management). Initially, we assessed each article by reading its title and abstract to determine if it met the inclusion criteria. Articles that only had an abstract available were subsequently excluded.

### Selection process

The eligibility assessment of the studies was performed by two reviewers independently. Titles and abstracts were screened to select only potentially eligible citations. Then, full-text publications of the selected citations that written in English language were retrieved and further screened for eligibility. In case of any discrepancies or disagreements, a consensus was reached through discussion and if needed resolved by a third researcher.

### Data collection process and data items

The data extraction process from individual studies was conducted by three reviewers independently using a structured Microsoft Excel spread sheet. The extracted data included the authors’ names, year of publication, study location/area, number of participants, study design, interventions, control groups, and primary outcome (VAS score during labor) and secondary outcomes (BP, HR, mode of delivery, Apgar score, complications…) of the included studies. All these details were carefully recorded and documented.

### Risk of bias assessment of the individual studies

The methodological quality of the included studies was assessed using the Cochrane Collaboration’s risk of bias assessment tool specifically designed for randomized controlled trials (RCTs). All the studies included in our systematic review underwent a critical appraisal and assessment of the risk of bias. We used the Cochrane risk of bias tool and appraised five domains: selection bias (random sequence generation and allocation concealment), performance bias (blinding of participants and personnel), detection bias (blinding of outcome assessment), and attrition bias (incomplete outcome data), and reporting bias (selective reporting), and other factors as described in Table 2 (annex 1). Each potential source of bias was graded as low, unclear, or high.

## Result

### Study selection

The comprehensive search strategy resulted in the identification of 38 articles from various electronic databases. After removing duplicates using the EndNote reference manager, 32 articles remained for screening. Among these, nine articles were ultimately included for critical appraisal, while the rest articles were excluded for various reasons. The results of the search strategy and the study selection process is illustrated in the PRISMA flow chart above (Fig. 1).

**Figure 1.**
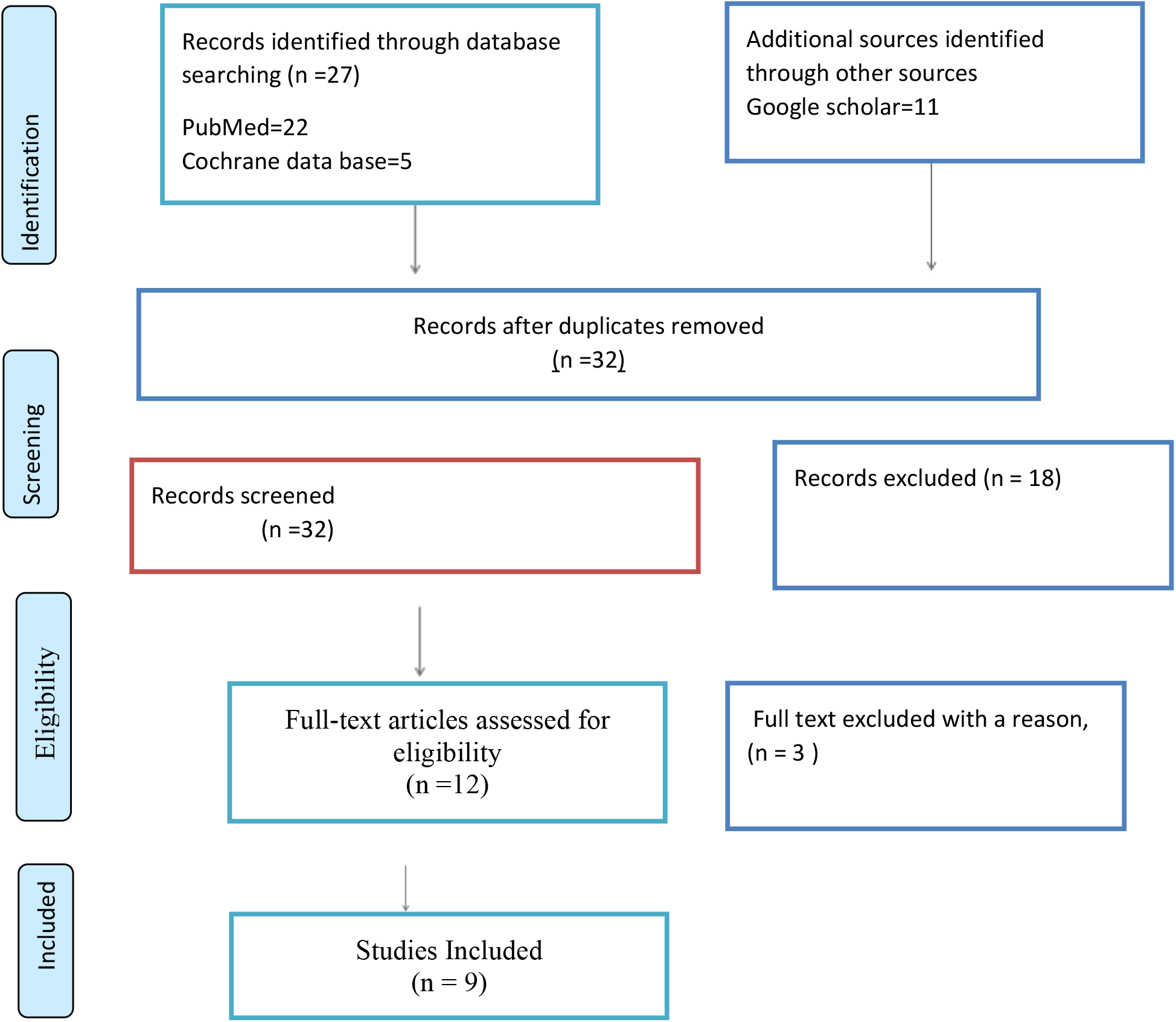
PRISMA Study flow diagram for selection, screening and inclusion of studies.

### Study and participants characteristics

All the nine studies included were randomized controlled trials (RCTs) comparing intrathecal Dexmedetomidine with opioids and placebo for single shot spinal or combined spinal epidural labor analgesia.

Of these nine studies were involved a comparative analysis of intrathecal dexmedetomidine with bupivacaine and fentanyl with bupivacaine for epidural labor analgesia whereas the rest two studies compared Dexmedetomidine with sufentanil (13-22). Of the nine studies included in the review, six were conducted in India, one in Nigeria, one in china and one in Kenya. A total of 911 participants were included in the nine studies. Additionally, a summary of the included studies in this review can be found in the table below (Table 1)

**Table 1.** Summary of included articles characteristics.

| *Authors year* | *Location* | *Study design* | *Participants* | *Intervention* | *Control* | *outcomes* | *ROB* |
| --- | --- | --- | --- | --- | --- | --- | --- |
| *S.Fynefac e-Ogan et al.2012* | Nigeria | RCT | 90 | Intrathecal Dex 2.5 µg | Fentanyl 2.5 µg | A single-shot intrathecal low-dose dexmedetomidine injection has the potential to significantly reduce pain during labor and delivery. Intrathecal 5µg and 10µg dexmedetomidine can cause prolonged sensory block with no effect on blood pressure or heart rate. | low |
| *Rathodetal et al.2021* | India | RCT | 120 | Intrathecal Dex 5µg | fentanyl 25 µg | single-shot-intrathecal labor analgesia using the combination of bupivacaine (2.5 mg), fentanyl (25 µg), and morphine (250 µg) or dexmedetomidine (5 µg) is a viable method of pain relief in labor and delivery | low |
| *Dilesh P.K et al.2014* | India | RCT | 112 | Intrathecal Dex 10 µg | Fentanyl 20µg | 10 µg dexmedetomidine is an attractive alternative to 20 µg Fentanyl as an intrathecal agent for labor analgesia. It provides acceptable quality of labor analgesia with increased duration of analgesia compared to fentanyl. | low |
| *Jain et al/2019* | India | RCT | 60 | Intrathecal Dex 5 µg | fentanyl 25 µg | In comparison to the addition of fentanyl, 5µg of dexmedetomidine for labor analgesia. Provides excellent and longer pain relief with minimal side effects and no delay in the progression of labor. | low |
| *Jain et al.2022* | India | R<br>C<br>T | 120 | Intrathecal<br>Dex 20 µg | fentanyl 15<br>µg | Intensity and duration of analgesia are better with intrathecal dexmedetomidine. But: Chances of vaginal delivery are higher with intrathecal fentanyl as an adjuvant. | low |
| *Gao et al. 2022* | Kenya | R<br>C<br>T | 91 | Intrathecal<br>Dex.<br>0.4µg/ml | Sufentanyl<br>0.4µg/ml | Dexmedetomidine outperforms opioid analgesic sufentanil in providing a prolonged analgesic effect as an epidural during labor. It also reduces the need for local anesthetics and has fewer side effects. | low |
| *Gehuili et al 2021* | China | R<br>C<br>T | 108 | intrathecal<br>5 µg Dex<br>and 5 µg<br>sufentanil<br>intrathecal | Saline | Intrathecal administration of 5 mg Dexmedetomidine or 5 mg sufentanil combined with 0.1% epidural ropivacaine and 0.2 mg/mL sufentanil was found to provide improved analgesia with quicker onset times and reduced need for local anesthetics compared to saline administration. | Low |
| *Verma et al.2023* | India | R<br>C<br>T | 90 | 10 µg<br>intrathecal<br>dexmedetomidine | 10 µg<br>fentanyl | Intrathecal dexmedetomidine 10 µg alone and a combination of 5 µg dexmedetomidine and 10 µg fentanyl provided adequate labor analgesia that was comparable to fentanyl alone | Low |
| Madishetti and Aasim's 2016 | India | R<br>C<br>T | 120 | 5 µg<br>intrathecal<br>dexmedetomidine | 10 µg<br>fentanyl | Adding 5 µg of intrathecal dexmedetomidine to 10 µg of fentanyl prolonged the duration of analgesia. | Low |

*Note: ROB=risk of bias, RCT=randomized control trial, DEX= Dexmedetomidine,*

**Table 2.** Risk of Bias Assessment for included studies.

| Studies | Selection bias |  | PB<br>(BPP) | D B<br>(B O) | AB<br>(IO) | RB<br>(SR) | OB |
| --- | --- | --- | --- | --- | --- | --- | --- |
|  | RSG | AC |  |  |  |  |  |
| *S.Fyneface-Ogan et al.2012* | + | + | + | + | + | + | - |
| *Dilesh P.K et al.2014* | + | + | ? | - | + | - | + |
| *Jain et al.2019* | + | + | - | ? | + | + | - |
| *Rathod et al.2021* | + | - | + | - | - | + | + |
| *Jain et al.2022* | + | + | ? | - | - | + | ? |
| *Gao et al. 2022* | + | + | + | + | - | + | - |
| *Gehuili et al 2021* | + | + | + | - | -+ | - | + |
| *Verma et al.2023* | + | + | + | + | + | + | + |
| **Madishetti and Aasim's 2016** | + | + | + | + | + | + | + |

Key1. ROB: 1-3=high risk (H), 4-7=low risk (L), 0=for NO, 1=for YES; 2=for cannot tell.
Key 2, RSG=random Sequence Generation. AC= allocation concealment. PB =Performance Bias (blinding participant & personal). DB=detection Bias (blinding of Outcomes). AB= attrition Bias (Incomplete outcomes). RB= reporting Bias (selective reporting).OB=Others Bias .ROB= Risk of Bias.

### Risk of bias assessment

The risk of bias is presented in table 2. (Annex) All the included studies are in low risk of bias with the score of 4-7. (Table 2)

## Discussion

According to this review, intrathecal dexmedetomidine provides a safe and effective adjunct both for single shot spinal and combined spinal epidural labor analgesia; providing potent analgesic efficacy with improved quality of the block and prolonged duration of analgesia with minimal risk of opioid related side effects for both mothers and neonates (13-22).

Dexmedetomidine provides prolonged analgesic duration. The prolonged analgesic effect of dexmedetomidine is due to its synergism with local anesthetics (13). This is consistent with its mode of action as an agonist of the α2-adrenergic receptor possessing both direct analgesic and potentiating local anesthetic effects. Dexmedetomidine ability to lessen the need for opioids has important therapeutic ramifications for preventing opioid-related side effects in both mothers and newborns. A meta-analysis by Paramasivan et al.2022 concludes that Intrathecal Dexmedetomidine provides prolonged duration of postoperative analgesia, decreased 24hr pain intensity, and reduced incidence of shivering compared to placebo. (4)

Intrathecal Dexmedetomidine also found as a safe and effective adjuvant when used as single-shot intrathecal labor analgesia combined with low dose bupivacaine. A study by Rathod et al. 2021 and Fyneface-Ogan et al. in 2012 demonstrated that single-shot intrathecal labor analgesia using a combination of low dose bupivacaine with dexmedetomidine vs with fentanyl for labor analgesia. (17, 18) Both studies favored dexmedetomidine over fentanyl and reported no adverse effect on neonatal outcome. This shows Dexmedetomidine is a promising simple, safe and effective spinal labor analgesia adjunct especially for resource limiting settings where epidural kit is unavailable. In addition to providing improved analgesia, Intrathecal dexmedetomidine administration had also decreased opioids related side effects like incidence of pruritus and shivering compared with when opioids are used as adjuvant in labor CSE (16, 22).

Dexmedetomidine can also be effective when used as adjunct for epidural labor analgesia through epidural route. Meta-analysis and Systematic review done on epidural Dexmedetomidine adjunct for labor analgesia by Li et al. 2021 concludes Dexmedetomidine when used as an adjuvant to epidural local anesthetics for labor analgesia has the potential to provide better analgesic effect than placebo, comparable labor pain control to opioid and has no definite adverse effect on parturient of fetus.(23)

Intrathecal Dexmedetomidine when added with dexamethasone it provide strong selective sensory block with prolonged duration of analgesia. (24) According to Ali et al. (2021), the use of Dexmedetomidine in conjunction with Ropivacaine-Dexamethasone was compared to Fentanyl as an adjunct for labor analgesia. The findings revealed that the combination of Ropivacaine-Dexamethasone and Dexmedetomidine provided a strong, selective sensory block with a prolonged duration of analgesia, a delayed S1 regression time, and limited maternal and fetal adverse effects when compared to the combination of Ropivacaine-Dexamethasone and Fentanyl. (24)

Studies favors the use of intrathecal Dexmedetomidine as adjunct to neuraxial labor analgesia. However, there is no recommended optimal dose of Dexmedetomidine. The studies included in this review used different doses of dexmedetomidine ranging from 2.5mcg to 20 mcg. (13-22) We recommend large sample size studies on optimal dose of Dexmedetomidine for intrathecal labour analgesia.

In addition, this review has several limitations. First of all, the small sample sizes of the studies that were part of the analysis made it difficult to identify uncommon negative effects. Secondly, there was no long-term monitoring for newborns. The available data, in spite of these drawbacks, favors the use of dexmedetomidine as a useful supplement to local anesthetics for labor epidural analgesia. It is a good substitute for opioid-based regimens because of its safety profile and effectiveness, especially in environments with low resources.

As conclusion, this review indicates that compared to opioids intrathecal administration of dexmedetomidine for labor analgesia provides potent analgesic efficacy, improving the duration, intensity, and quality of the block, reducing the need for opioids and minimizing opioid related adverse effects for both mothers and neonates.

## Data Availability

All relevant data are within the manuscript and its Supporting Information files

## Declaration

**Ethics approval and consent to participate**: N/A

**Publication consent:** N/A

**Competing interest**: No

**Funding source**: N/A

## Author’s contributions

The corresponding author, Adanech S Legasse and Fikadu Tilahun contributes in the study by conceptualization, methodology, Data extraction, writing and editing the review. Bekele Buli, Yesuf Ahmed, Addisu Mossie, Aschalew Besha and Selam Tamiru have made a significant contribution in conceptualization, methodology, data extraction and writing draft.

## Declaration of generative AI and AI-assisted technologies in the writing process

During the preparation of this work the author(s) used paperpal in order to rewrite and editing. After using this tool/service, the author(s) reviewed and edited the content as needed and take(s) full responsibility for the content of the publication.

## Annexes Annex 1. Risk of bias assessment table

